# Longitudinal assessment of lung function in Swiss childhood cancer survivors

**DOI:** 10.1101/2022.03.16.22272460

**Authors:** Rahel Kasteler, Maria Otth, Florian S Halbeisen, Luzius Mader, Florian Singer, Jochen Rössler, Nicolas X von der Weid, Marc Ansari, Claudia E Kuehni

## Abstract

**Rationale:** Childhood cancer survivors (CCSs) are at increased risk for pulmonary morbidity due to exposure to lung-toxic treatments, including specific chemotherapies, radiotherapy, and surgery. Longitudinal data on lung function, with information on how outcomes change over time, in CCSs are scarce.

**Objectives:** To investigate lung function trajectories in childhood cancer survivors over time and investigate the association with lung-toxic treatment.

**Methods:** This retrospective, multi-center cohort study included CCSs, who were diagnosed between 1990 and 2013 in Switzerland and had been exposed to lung-toxic chemotherapeutics or thoracic radiotherapy. Pulmonary function tests (PFTs) were obtained from hospital charts. We assessed quality of PFTs systematically and calculated z-scores and percentage predicted of forced expiratory volume in first second (FEV1), forced vital capacity (FVC), FEV1/FVC ratio, total lung capacity (TLC) and diffusion capacity for carbon-monoxide (DLCO) based on recommended reference equations. We described lung function over time and determined risk factors for change in FEV1 and FVC using multivariable linear regression.

**Results:** We included 790 PFTs from 183 CCSs, with a median age of 12 years (IQR 7 – 14) at diagnosis. Common diagnoses were lymphoma (55%), leukemia (11%) and CNS tumors (12%). Median follow-up time was 5.5 years. Half (49%) of CCSs had at least one abnormal pulmonary function parameter, with restrictive impairment being common (22%). Trajectories of FEV1 and FVC started at z-scores of -1.5 at diagnosis and remained low throughout follow-up. CCSs treated with thoracic surgery started particularly low, with an FEV1 of -1.08 z-scores (−2.02 to -0.15) and an FVC of -1.42 z-scores (−2.27 to -0.57) compared to those without surgery. In CCS exposed to lung-toxic chemotherapeutics FEV1 z-scores increased slightly over time (0.12 per year; 95%CI 0.02 - 0.21).

**Conclusion:** The large proportion of CCSs with reduced lung function identified in this study underlines the need for more research and long-term surveillance of this vulnerable population.

## Introduction

Childhood Cancer Survivors have an increased risk of pulmonary mortality and morbidity compared to their peers (1-4). Lung-toxic treatments, currently inevitable to treat cancer, can cause irreversible damage to the lung. Established risk factors are the chemotherapeutics bleomycin, busulfan and nitrosureas (lomustine and carmustine), thoracic radiotherapy, and thoracic surgery (5, 6). Survivors report more pulmonary symptoms, such as exercise intolerance or chronic cough, and suffer more often from recurrent pneumonia, lung fibrosis or emphysema than siblings (3, 4, 7). They are also at higher risk for hospitalization and mortality from pulmonary causes (8-10). Cancer treatment induced respiratory diseases may remain subclinical while lung function impairment progresses and ultimately leads to premature respiratory morbidity and mortality (11). Pulmonary function tests (PFTs) may to detect pulmonary dysfunction at an earlier, asymptomatic stage and are therefore recommended in long-term follow-up guidelines for survivors (12-14).

Only one study reported longitudinal data on pulmonary function in survivors of all cancer types (11). Other studies focused on specific cancer entities or treatments groups, such as embryonal brain tumors, Hodgkin lymphoma, whole lung irradiation or hematopoietic stem cell transplantation (15-24). Most publications described single center studies, only one was a multicenter study of survivors enrolled in a specific treatment protocol for embryonal brain tumors (17). We could not find data on an unselected, national sample of survivors. Interpretation of several previous studies is constrained as lung function was expressed as percent predicted, which is subject to sex, age, height and ethnicity bias - especially in pediatric populations (25). Flawed transformation of lung function indices hampers comparison across age groups and between studies (26). The Global Lung Initiative (GLI) published international sex-, age-, and height-adjusted prediction equations for spirometry (2012), plethysmography (2021), and diffusion capacity of carbon monoxide (DLCO, 2017) that cover patients aged 3-95 years and many ethnic groups (27-29).

In this national study, we describe longitudinal data on pulmonary function in Swiss survivors diagnosed 1990-2013 and exposed to lung-toxic treatments. We used the GLI equations and z-scores to describe longitudinal trajectories for forced expiratory volume in the first second (FEV1) and forced vital capacity (FVC) and determined factors associated with deterioration or improvement of lung function over time.

## Methods

### Study population

This study is nested within the Swiss Childhood Cancer Registry (SCCR), a national, population-based registry of all children and adolescents who were diagnosed with leukemia, lymphoma, central nervous system tumors, malignant solid tumors, or Langerhans cell histiocytosis prior to the age of 21 years and were living in Switzerland at time of diagnosis (30). The SCCR collects information on diagnosis, classified according to the International Classification of Childhood Cancer, third edition (ICCC-3) (31), cancer treatments, and demographic information such as date of birth and sex. Ethics approval was granted by the Ethics Committee of the Canton of Bern to the SCCR (KEK-BE: 166/2014 and 2021-01462).

### Inclusion criteria

We included all survivors from the SCCR who had been diagnosed between 1990 and 2013 in any of the nine Swiss pediatric oncology group centers before age 16 years, had survived two years or more, and had been exposed to at least one lung-toxic chemotherapeutic agent or thoracic radiotherapy. We excluded survivors whose medical records were not available or did not contain pulmonary function tests (PFT) following the cancer diagnosis, and those who were aged ≤ 6 years at time of the study, because of limited quality of PFTs. We defined exposure to lung-toxic chemotherapeutics as treatment with busulfan, bleomycin, and/or nitrosoureas (lomustine, carmustine), following international long-term follow-up care guidelines (13, 14, 32). Radiotherapy was considered as lung-toxic when administered as total body irradiation (TBI) or involving the mantle-field, chest, lungs, mediastinum or thoracic spine, including craniospinal irradiation (CSI) (13, 14, 32). We recorded information on thoracic surgery performed by thoracotomy or thoracoscopy for tumor/metastasis resection (wedge, lobe, whole lung), rib resection, laminectomy, bone biopsy, en bloc resection of rib, lung tissue, and/or diaphragm. Of all lung-toxic exposures we collected the date of first administration.

### Medical records review and pulmonary function measurements

We extracted information on date of birth, sex, diagnosis, date of diagnosis, and treatment protocol from the SCCR. We collected results of spirometry, body plethysmography and DLCO, and information on treatment exposure from electronic and paper-based medical records until December 31th 2015. We searched the archives of the Pediatric Oncology and Pediatric Pulmonology clinics where the children had been treated. Pediatric oncologists refer survivors to PFT laboratories in the tertiary care pediatric pulmonology departments on site, which conduct PFTs according to ATS/ERS guidelines (33-35). We excluded PFTs performed more than 10 years after diagnosis. These were available only for few survivors and including them would have introduced selection bias, because those with poorer lung function may have had extended follow-up.

From the PFT results we extracted test date, height and weight, and the following outcome measures: FVC [l], FEV1 [l], total lung capacity (TLC [l]), and DLCO [mmol/min/kPa]. We calculated BMI [kg/m^2^] and BMI z-scores using national reference data (36). We performed a quality check of PFTs according to ATS/ERS guidelines (33-35). We always used the measurement prior to bronchodilatation because post-bronchodilation tests had been done rarely. We converted all outcomes into sex-, height-, and age-adjusted z-scores and percentage predicted using the Global Lung Initiative (GLI) 2012 reference values for FEV1, FVC, FEV1/FVC, TLC and DLCO (27-29) (26, 33). We defined lower limits of normality (LLN) as z-scores of less than -1.645 (37).

To ease comparison with previous reports (11, 38, 39) we also categorized outcomes into the following binary categories: FVC and FEV1 were defined as abnormal when they were of <80% predicted, and the FEV1/FVC ratio when it was <0.70 (40). DLCO and TLC were defined as abnormal when they were <75% of predicted according to the Common Terminology Criteria for Adverse Events version 3.0 (CTCAE v3.0) (41). These cut-offs are comparable with previous reports, though not exactly in accordance with ATS/ERS interpretative strategies (42).

We grouped pulmonary dysfunction into three outcomes: restrictive impairment (TLC z-score <-1. 645 or TLC < 75% and FEV1 ≥ 80% predicted), obstructive impairment (FEV1/FVC <0.70 and FEV1 <80% of predicted or FEV1/FVC <0.70 and FEV1 z-score <-1.645), and diffusion capacity impairment (DLCO <75% predicted or z-score <-1.645). For this categorization we used the last PFT result in those who had more than one test and the available result in those with one test only.

### Statistical analysis

We used medians and interquartile ranges (IQR) to describe the data. We primarily described the PFT results in the last test of each survivor, independent of whether all PFT parameters have been assessed at this time point. In a sensitivity analysis we included only survivors, in whom all PFT parameters had been measured in the last test. In survivors with at least two PFTs, we compared z-scores for FVC, FEV1, and FEV1/FVC ratio between first and last test using Wilcoxon rank-sum test. We plotted trajectories of z-scores for FEV1 and FVC by fitting a loess curve (locally weighted smoothing) and the respective 95% confidence interval (CI). We evaluated the association between potential risk factors and FEV1 and FVC trajectories using multivariable linear mixed effects regression models with random intercept and random slope, to account for repeated measurements within patients. Potential risk factors were time since first exposure to a lung-toxic treatment, sex, BMI z-score, age at diagnosis, cancer diagnosis, radiotherapy, lung-toxic chemotherapy, HSCT, and thoracic surgery. We included time since first exposure to a lung-toxic treatment as a linear term and did not take autocorrelation into account. We also evaluated potential effect modification of FEV1 or FVC slopes over time by including interaction terms between risk factors and time since cancer diagnosis. We performed likelihood ratio tests to evaluate associations between risk factors and changes in FEV1 or FVC. Age at diagnosis was moderately associated with time since diagnosis. We therefore included this variable in an additional sensitivity analysis as a categorical variable (0-4 years, 5-13 years, >13 years). All analyses were performed using Stata (Version 16, Stata Corporation, Austin, Texas) or R 3.1.2 (www.r-project.org). Linear mixed models were performed using the R-package lmer.

## Results

### Characteristics of the study population

Of 2989 survivors registered in the SCCR, 372 (12%) had been exposed to lung-toxic treatment, had traceable medical records, and were at least six years old at data collection. Records of half of these patients (183, 49%) included PFTs, which had been performed after the lung-toxic treatment and within 10 years from diagnosis **(Supplementary Figure E1)**. Median age at diagnosis was 12.4 years (interquartile range, IQR 7-14), 58% were male, the most common cancer diagnosis was lymphoma (55%), and 25% had suffered a relapse (**Table 1**). Eighty-seven percent had received thoracic radiotherapy, 39% lung-toxic chemotherapy, 23% HSCT, and 10% thoracic surgery.

**Table 1.**
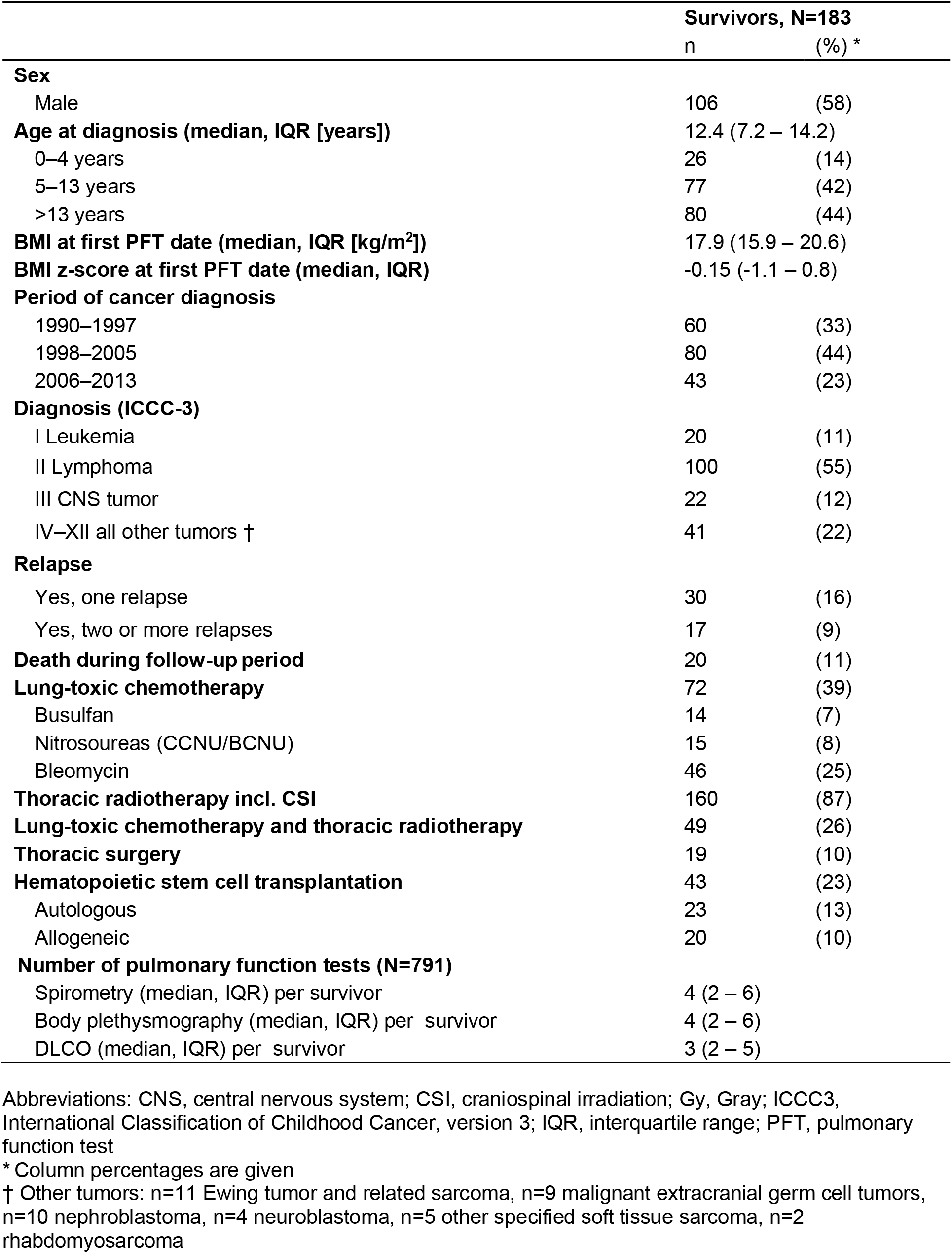
Characteristics of Swiss childhood cancer survivors exposed to lung-toxic therapies between 1990 and 2013 (N=183)

|  | Survivors, N=183 |  |
| --- | --- | --- |
|  | n | (%) * |
| **Sex** |  |  |
| Male | 106 | (58) |
| **Age at diagnosis (median, IQR [years])** | 12.4 (7.2 – 14.2) |  |
| 0–4 years | 26 | (14) |
| 5–13 years | 77 | (42) |
| >13 years | 80 | (44) |
| **BMI at first PFT date (median, IQR [kg/m<sup>2</sup>])** | 17.9 (15.9 – 20.6) |  |
| **BMI z-score at first PFT date (median, IQR)** | -0.15 (-1.1 – 0.8) |  |
| **Period of cancer diagnosis** |  |  |
| 1990–1997 | 60 | (33) |
| 1998–2005 | 80 | (44) |
| 2006–2013 | 43 | (23) |
| **Diagnosis (ICCC-3)** |  |  |
| I Leukemia | 20 | (11) |
| II Lymphoma | 100 | (55) |
| III CNS tumor | 22 | (12) |
| IV–XII all other tumors † | 41 | (22) |
| **Relapse** |  |  |
| Yes, one relapse | 30 | (16) |
| Yes, two or more relapses | 17 | (9) |
| **Death during follow-up period** | 20 | (11) |
| **Lung-toxic chemotherapy** | 72 | (39) |
| Busulfan | 14 | (7) |
| Nitrosoureas (CCNU/BCNU) | 15 | (8) |
| Bleomycin | 46 | (25) |
| **Thoracic radiotherapy incl. CSI** | 160 | (87) |
| **Lung-toxic chemotherapy and thoracic radiotherapy** | 49 | (26) |
| **Thoracic surgery** | 19 | (10) |
| **Hematopoietic stem cell transplantation** | 43 | (23) |
| Autologous | 23 | (13) |
| Allogeneic | 20 | (10) |
| **Number of pulmonary function tests (N=791)** |  |  |
| Spirometry (median, IQR) per survivor | 4 (2 – 6) |  |
| Body plethysmography (median, IQR) per survivor | 4 (2 – 6) |  |
| DLCO (median, IQR) per survivor | 3 (2 – 5) |  |

Abbreviations: CNS, central nervous system; CSI, craniospinal irradiation; Gy, Gray; ICCC3, International Classification of Childhood Cancer, version 3; IQR, interquartile range; PFT, pulmonary function test
\* Column percentages are given
† Other tumors: n=11 Ewing tumor and related sarcoma, n=9 malignant extracranial germ cell tumors, n=10 nephroblastoma, n=4 neuroblastoma, n=5 other specified soft tissue sarcoma, n=2 rhabdomyosarcoma

### Lung function

Records of the 183 participants contained 790 PFTs done within the follow-up period, with a median of four tests per participant for spirometry and body plethysmography, and three measurements of DLCO (**Table 1**). Among the 183 participants, 32 (17%) had PFTs done at one occasion only, and 151 (83%) had repeated tests. The cross-sectional analysis included the last available test from all 183 participants, which included 180 measurements of FEV1, 178 of FVC, 176 of FEV1/FVC, 166 TLC values and 103 results from DLCO (**Table 2**). The median follow-up time from diagnosis to the last PFT was 5.5 years (IQR 2.6 – 7.7 years). FEV1 and FVC z-scores were below -1.645 in 33% and 38% of participants respectively. Only two patients (1%) had a FEV1/FVC ratio below 0.7 and an FEV1 z-score below -1.645, meeting the GOLD-criteria for obstructive disease **(Table 2)**. TLC z-scores were below -1.645 in 22% of participants, indicating restrictive lung disease. Diffusion capacity was reduced in 25%. In half of the participants (49%), at least one outcome (FEV1, FVC, TLC or DLCO) was abnormal (z-score <-1.645) and 31% fulfilled the criteria of either obstructive disease, restrictive disease or diffusion capacity impairment. When using percentage predicted instead of z-scores, the proportion of survivors with values below the LLN was similar for FEV1 (31%) and FVC (36%), and slightly lower for DLCO (18%) **(Table 2)**. Our sensitivity analysis, based on the last test when all five outcomes were measured at the same time, included 128 survivors (**Supplementary Table E1**). Results were similar to the main analysis.

**Table 2.** Pulmonary function in childhood cancer survivors exposed to lung-toxic therapies, described as z-scores and percentage predicted (cross-sectional analysis based on last available test, N=183)

|  | Survivors |  |  |
| --- | --- | --- | --- |
|  | N* | % | median (IQR) |
| **Time from diagnosis to last test [years]** | 183 |  | 5.5 (2.6 – 7.7) |
| **Age at last PFT (median, IQR [years])** | 183 |  | 16.7 (12.6 – 19.3) |
| **FEV1** |  |  |  |
| FEV1 z-score | 180 |  | -1.1 (-1.9 – -0.3) |
| FEV1 z-score <-1.645 | 59 | 33 | -2.4 (-3.2 – -1.9) |
| FEV1% predicted | 180 |  | 87.5 (77.4 – 96.7) |
| FEV1 <80%predicted | 56 | 31 | 70.6 (61.3 – 76.9) |
| **FVC** |  |  |  |
| FVC z-score | 178 |  | -1.3 (-2.2 – -0.3) |
| FVC z-score <-1.645 | 68 | 38 | -2.5 (-3.2 – -2.1) |
| FVC% predicted | 178 |  | 84.2 (74.1 – 96.1) |
| FVC <80% predicted | 64 | 36 | 70.6 (62.3 – 75.4) |
| **FEV1/FVC** |  |  |  |
| FEV1/FVC z-score | 176 |  | 0.2 (-0.4 – 1.1) |
| FEV1/FVC z-score <-1.645 | 4 | 2 | -2.1 (-2.4 – -1.8) |
| FEV1/FVC% predicted | 176 |  | 101 (96.9 – 107.3) |
| FEV1/FVC <0.7 | 2 | 1 | 0.68, 0.68 |
| FEV1/FVC <0.7 and FEV1<80%predicted | 1 | <1 | n.a. |
| FEV1/FVC <0.7 and FEV1 z-score <-1.645 | 2 | 1 | n.a. |
| **TLC** |  |  |  |
| TLC z-score | 166 |  | 0.7 (-1.47 – -0.03) |
| TLC z-score <-1.645 | 37 | 22 | -2.1 (-2.9 – -1.8) |
| TLC % predicted | 166 |  | 91.4 (81.5 – 100.5) |
| TLC <75% predicted | 24 | 15 | 67 (61.1 – 72.15) |
| TLC <75% predicted and FEV1 ≥ 80% predicted | 2 | 1 | n.a. |
| **DLCO** |  |  |  |
| DLCO z-score | 103 |  | -0.7 (-1.6 – 0.5) |
| DLCO z-score <-1.645 | 25 | 25 | -2.6 (-3.4 – -1.8) |
| DLCO% predicted | 103 |  | 89.7 (78.6 – 106.8) |
| DLCO <75% predicted<sup>c</sup> | 18 | 18 | 60.1 (53.5 – 69.2) |
| **Any impairment** |  |  |  |
| FEV1, FVC, TLC or DLCO z-score < -1.645 | 89 | 49 | n.a. |
| Obstructive †, restrictive ‡, or diffusion capacity impairment § | 56 | 31 | n.a. |

Abbreviations: IQR, interquartile range; PFT, pulmonary function test
\* As not every last test included spirometry, body plethysmography and DLCO, the total number of CCSs per test parameter does not sum up to 183
† Obstructive = FEV1/FVC<0.7 and FEV1 z-score <-1.645
‡ Restrictive = TLC z-score <-1.645
§ Diffusion capacity impairment = DLCO z-score <-1.645

We then compared changes between first and last PFT in the 151 participants with repeated tests (**Table 3**). The median time from diagnosis to the first PFT was 0.8 years (IQR 0.3 – 1.6) and the median time between the first and the last test was 4.2 years (2.5-6.2). None of the outcomes changed over time and the mean difference between the results of the first and the last test was small. This is also evident when visualizing the longitudinal trajectories of FEV1 and FVC, which started at z-scores of around -1.5 and did not improve or deteriorate during the observed period (**Figure 1a and Figure 1b**).

**Table 3.** Change in pulmonary function outcome parameters (FEV1, FVC, FEV1/FVC) between the first and the last test in childhood cancer survivors exposed to pulmotoxic treatment, described as z-scores and as percentage predicted (N=151)

|  | Survivors |  | Mean difference * | p-value † |
| --- | --- | --- | --- | --- |
|  | First Test | Last Test |  |  |
|  | median (IQR) | median (IQR) | median (IQR) |  |
| **Time from diagnosis to test (median, IQR [years])** | 0.8 (0.3 – 1.6) | 5.5 (2.6 – 7.7) | 4.2 (2.5-6.2) | NA |
| **Age at PFT (median, IQR [years])** | 13.7 (8.5 – 15.4) | 17.5 (13.5 – 19.7) | NA | NA |
| **FEV1 (n=147) ‡** |  |  |  |  |
| FEV1 z-score | -0.9 (-2.1 - -0.2) | -1.1 (-1.9 - -0.06) | 0.06 (-0.7 – 0.7) | 0.943 |
| FEV1% predicted | 88.8 (74.3 – 98.0) | 87.5 (77.5 – 99.3) | 1.8 (-7.9 – 7.9) | 0.962 |
| **FVC (n=149) ‡** |  |  |  |  |
| FVC z-score | -1.3 (-2.1 - -0.1) | -1.3 (-2.1 - -0.3) | 0.04 (-0.7 – 0.7) | 0.917 |
| FVC% predicted | 84.6 (74.8 – 98.3) | 84.7 (74.9 – 96.8) | 0.4 (-6.8 – 8.8) | 0.853 |
| **FEV1/FVC (n=145) ‡** |  |  |  |  |
| FEV1/FVC z-score | 0.6 (-0.5 – 1.4) | 0.2 (-0.4 – 1.0) | -0.2 (-0.9 – 0.5) | 0.146 |
| FEV1/FVC% predicted | 103.6 (96.2 – 108.6) | 101.4 (96.7 – 106.6) | -1.1 (-5.5 – 3.8) | 0.240 |

Abbreviations: IQR, interquartile range; PFT, pulmonary function test
\* Difference in time or z-score between first and last pulmonary function test per survivor taken into account
† Wilcoxon rank sum test
‡ Total number per parameter differs, as not all survivors had all three measurements at first and last test

**Figure 1:**
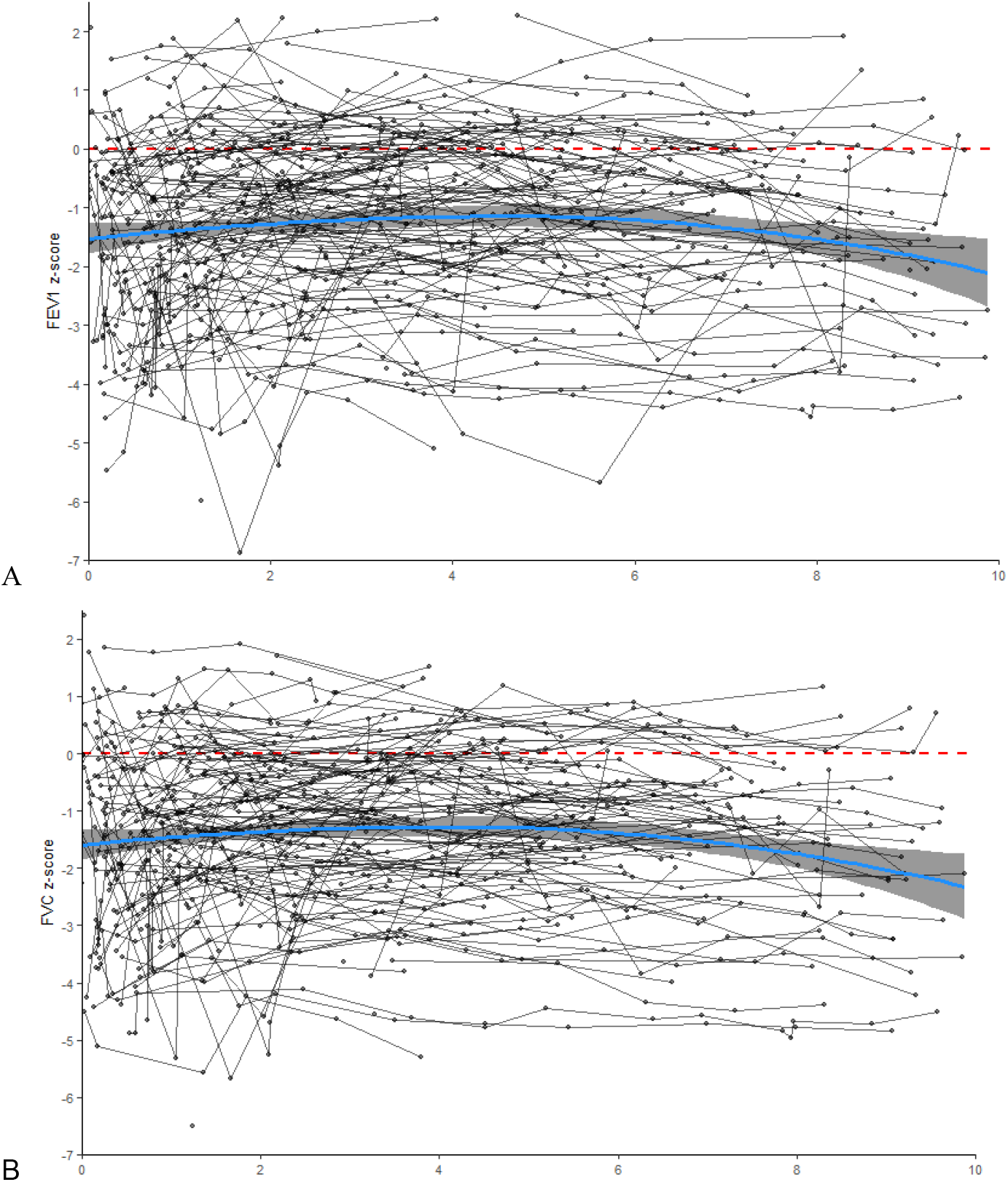
Longitudinal changes in FEV1 z-score (Panel A) and FVC z-score (Panel B) of childhood cancer survivors compared to Global Lung Function Initiative 2012 reference. Time zero corresponds to the date of diagnosis. The loess curve (blue line) shows the best fitted line by taking all data points into account at each time point on the x-line. The shaded band corresponds to the 95% confidence interval. Dots represent single test results, which are combined with a line and show the individual trajectories of each patient included in the study. The dashed red line represents the mean z-score of the normal population.

### Risk factors

In a multivariable linear mixed regression model, we assessed whether sex, BMI, cancer diagnosis and lung-toxic treatments were associated with the baseline of FEV1 and FVC (the intercept) and their change over time (the slope). The intercept was -1.07 z-scores for FEV1 (95%CI -2.18 – 0.03) and -1.57 z-scores for FVC (95%CI -2.67 – -0.47) (**Table 4; Table 5**). In this model, the intercept corresponds to the estimated lung function of the reference patient at diagnosis, not to the z-score at the time of the PFT. For further explanation of the model see **Supplementary Explanation E1**. Treatments were associated with baseline lung function. For FEV1, we found that children treated with thoracic surgery had a significantly lower intercept (−1.08 z-scores; 95%CI -2.02 – -0.15, **Table 4**) compared to those without. Adding this to the intercept of the reference patient, this means that survivors treated with thoracic surgery started at a FEV1 of -2.15 z-scores below reference values. Children exposed to lung-toxic chemotherapeutics tended to have lower FEV1 z-scores at diagnosis, which improved over time (0.12 per year; 95%CI 0.02 – 0.21, **Table 4**). HSCT was not significantly associated with baseline values but there was a trend for decreasing lung function over time (p=0.057). For FVC, thoracic surgery was again associated with lower z-scores at time of diagnosis (−1.42, 95%CI -2.27 – -0.57; **Table 5**) but no change over time. We also found a marginal association with type of tumor. FEV1 z-scores tended to be lower in children with bone tumors compared to those with lymphoma and did not change over time. In children with germ cell tumors, FEV1 and FVC tended to be higher at diagnosis, but decreased over time compared to those with lymphoma. Children with CNS and renal tumors had a decreasing FVC over time (**Table 4 and 5)**.

**Table 4.** FEV1 z-scores in childhood cancer survivors exposed to pulmotoxic treatment compared to GLI 2012 reference values; multivariable linear mixed regression adjusted for all covariates in the model (180 subjects, 760 tests)

|  | Estimate | 95% CI |  | p-value * |
| --- | --- | --- | --- | --- |
| **Intercept (FEV1 z-score at time of diagnosis of “reference patient” †)** | -1.07 | -2.18 | 0.03 |  |
| **Sex (ref. female)** | 0.16 | -0.30 | 0.61 | 0.502 |
| **BMI (ref. Healthy: z-score $\geq -2$, $\leq 1$)** |  |  |  | 0.419 |
| Underweight (z-score $< -2$) | -0.31 | -0.75 | 0.12 | |
| Overweight (z-score $\geq 1$, $\leq 2$) | 0.15 | -0.33 | 0.62 | |
| Obese (z-score $> 2$) | 0.23 | -0.40 | 0.86 | |
| **Cancer diagnosis (ref. lymphoma)** |  |  |  | 0.053 |
| CNS tumor | 0.08 | -0.63 | 0.79 |  |
| Bone tumors | -1.23 | -2.46 | -0.0003 |  |
| Germ cell tumors | 1.46 | 0.15 | 2.79 |  |
| Leukemia | -0.70 | -1.89 | 0.49 |  |
| Neuroblastoma | -1.27 | -3.12 | 0.58 |  |
| Renal tumors | 0.35 | -0.76 | 1.48 |  |
| Soft tissue sarcoma | -0.88 | -2.06 | 0.30 |  |
| **Thoracic radiotherapy (ref. no)** | 0.16 | -0.83 | 1.16 | 0.756 |
| **Chemotherapy (ref. no)** | -0.42 | -0.96 | 0.10 | 0.114 |
| **HSCT (ref. no)** | 0.28 | -0.53 | 1.09 | 0.479 |
| **Thoracic surgery (ref. no)** | -1.08 | -2.02 | -0.15 | 0.024 |
| **Change of FEV1 z-score per year** |  |  |  |  |
| **Time since diagnosis (continuous, per year)** | 0.09 | -0.12 | 0.31 |  |
| **Interaction male sex** | -0.07 | -0.15 | 0.01 | 0.106 |
| **Interaction BMI (ref. healthy: z-score $\geq -2$, $\leq 1$)** |  |  |  | 0.976 |
| Underweight (z-score $< -2$) | 0.02 | -0.18 | 0.13 | |
| Overweight (z-score $\geq 1$, $\leq 2$) | -0.02 | -0.11 | 0.07 | |
| Obese (z-score $> 2$) | -0.002 | -0.13 | 0.13 | |
| **Interaction cancer diagnosis (ref. lymphoma)** |  |  |  | 0.027 |
| CNS tumor | -0.12 | -0.25 | 0.01 |  |
| Bone tumors | 0.13 | -0.08 | 0.34 |  |
| Germ cell tumor | -0.28 | -0.51 | -0.05 |  |
| Leukemia | 0.13 | -0.07 | 0.34 |  |
| Neuroblastoma | 0.31 | -0.02 | 0.64 |  |
| Renal tumors | -0.09 | -0.26 | 0.08 |  |
| Soft tissue sarcoma | 0.004 | -0.20 | 0.21 |  |
| **Interaction thoracic radiotherapy (ref. no)** | -0.05 | -0.25 | 0.15 | 0.625 |
| **Interaction chemotherapy (ref. no)** | 0.12 | 0.02 | 0.21 | 0.016 |
| **Interaction HSCT (ref. no)** | -0.13 | -0.27 | 0.003 | 0.057 |
| **Interaction thoracic surgery (ref. no)** | -0.04 | -0.20 | 0.12 | 0.625 |

Abbreviations: CNS, central nervous system; HSCT, hematopoietic stem cell transplantation
\* Likelihood ratio test p-value indicating whether the characteristic explains differences in FEV<sub>1</sub> within the study population.
† Reference patient corresponds to a female person, at diagnosis, diagnosed with lymphoma, and not exposed to thoracic radiotherapy, chemotherapy, HSCT, and thoracic surgery.

**Table 5.**
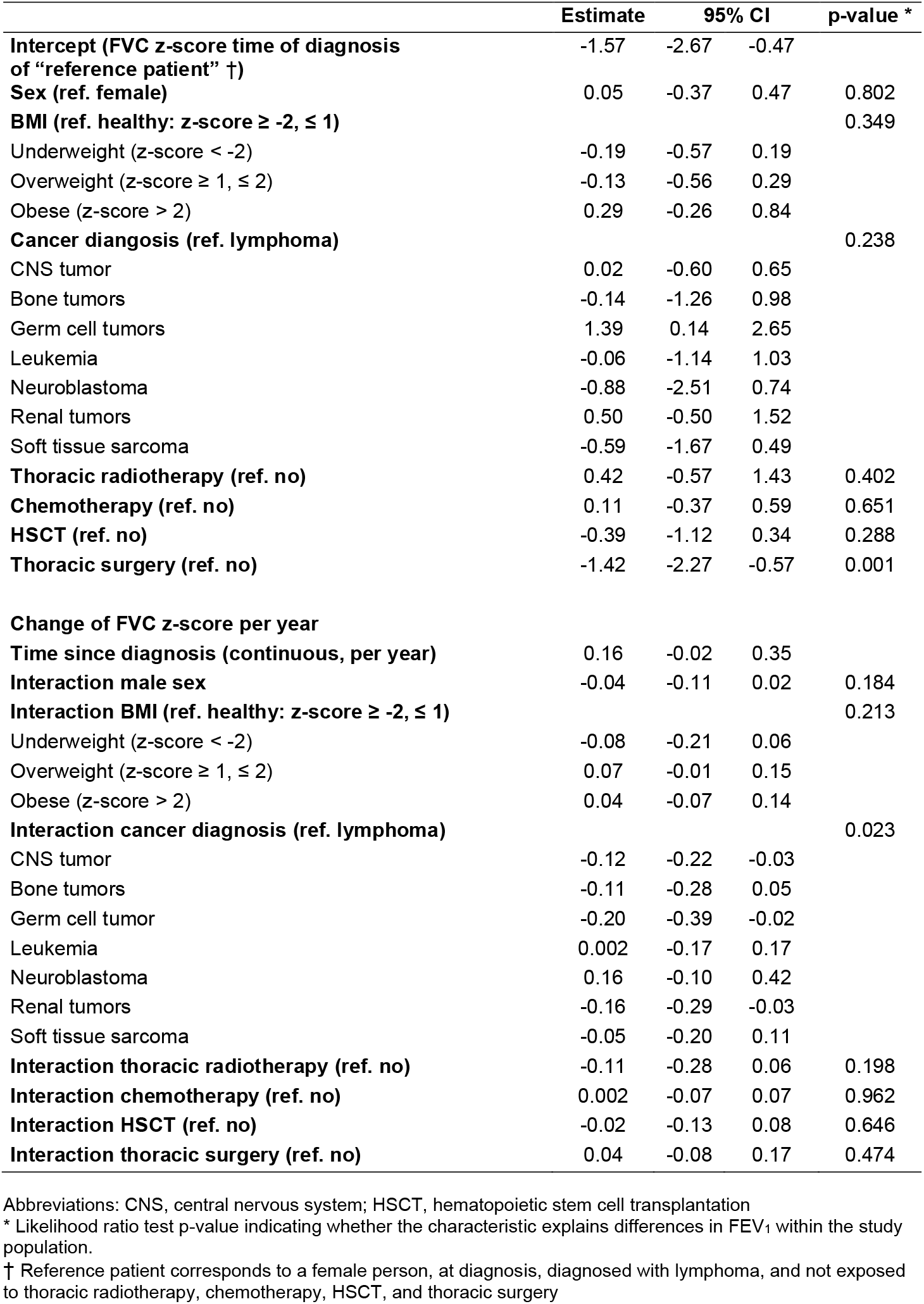
FVC z-scores in childhood cancer survivors exposed to pulmotoxic treatment compared to GLI 2012 reference values; multivariable linear mixed regression adjusted for all covariates in the model (178 subjects, 758 tests)

|  | Estimate | 95% CI |  | p-value * |
| --- | --- | --- | --- | --- |
| **Intercept (FVC z-score time of diagnosis of “reference patient” †)** | -1.57 | -2.67 | -0.47 |  |
| **Sex (ref. female)** | 0.05 | -0.37 | 0.47 | 0.802 |
| **BMI (ref. healthy: z-score $\geq -2$, $\leq 1$)** |  |  |  | 0.349 |
| Underweight (z-score $< -2$) | -0.19 | -0.57 | 0.19 | |
| Overweight (z-score $\geq 1$, $\leq 2$) | -0.13 | -0.56 | 0.29 | |
| Obese (z-score $> 2$) | 0.29 | -0.26 | 0.84 | |
| **Cancer diagnosis (ref. lymphoma)** |  |  |  | 0.238 |
| CNS tumor | 0.02 | -0.60 | 0.65 |  |
| Bone tumors | -0.14 | -1.26 | 0.98 |  |
| Germ cell tumors | 1.39 | 0.14 | 2.65 |  |
| Leukemia | -0.06 | -1.14 | 1.03 |  |
| Neuroblastoma | -0.88 | -2.51 | 0.74 |  |
| Renal tumors | 0.50 | -0.50 | 1.52 |  |
| Soft tissue sarcoma | -0.59 | -1.67 | 0.49 |  |
| **Thoracic radiotherapy (ref. no)** | 0.42 | -0.57 | 1.43 | 0.402 |
| **Chemotherapy (ref. no)** | 0.11 | -0.37 | 0.59 | 0.651 |
| **HSCT (ref. no)** | -0.39 | -1.12 | 0.34 | 0.288 |
| **Thoracic surgery (ref. no)** | -1.42 | -2.27 | -0.57 | 0.001 |
| **Change of FVC z-score per year** |  |  |  |  |
| **Time since diagnosis (continuous, per year)** | 0.16 | -0.02 | 0.35 |  |
| **Interaction male sex** | -0.04 | -0.11 | 0.02 | 0.184 |
| **Interaction BMI (ref. healthy: z-score $\geq -2$, $\leq 1$)** |  |  |  | 0.213 |
| Underweight (z-score $< -2$) | -0.08 | -0.21 | 0.06 | |
| Overweight (z-score $\geq 1$, $\leq 2$) | 0.07 | -0.01 | 0.15 | |
| Obese (z-score $> 2$) | 0.04 | -0.07 | 0.14 | |
| **Interaction cancer diagnosis (ref. lymphoma)** |  |  |  | 0.023 |
| CNS tumor | -0.12 | -0.22 | -0.03 |  |
| Bone tumors | -0.11 | -0.28 | 0.05 |  |
| Germ cell tumor | -0.20 | -0.39 | -0.02 |  |
| Leukemia | 0.002 | -0.17 | 0.17 |  |
| Neuroblastoma | 0.16 | -0.10 | 0.42 |  |
| Renal tumors | -0.16 | -0.29 | -0.03 |  |
| Soft tissue sarcoma | -0.05 | -0.20 | 0.11 |  |
| **Interaction thoracic radiotherapy (ref. no)** | -0.11 | -0.28 | 0.06 | 0.198 |
| **Interaction chemotherapy (ref. no)** | 0.002 | -0.07 | 0.07 | 0.962 |
| **Interaction HSCT (ref. no)** | -0.02 | -0.13 | 0.08 | 0.646 |
| **Interaction thoracic surgery (ref. no)** | 0.04 | -0.08 | 0.17 | 0.474 |

Abbreviations: CNS, central nervous system; HSCT, hematopoietic stem cell transplantation
\* Likelihood ratio test p-value indicating whether the characteristic explains differences in FEV<sub>1</sub> within the study population.
† Reference patient corresponds to a female person, at diagnosis, diagnosed with lymphoma, and not exposed to thoracic radiotherapy, chemotherapy, HSCT, and thoracic surgery

In the sensitivity analysis that included age at diagnosis in the regression model, the age group of 13 years and older at diagnosis tended to have a lower FEV1 z-score compared to those diagnosed at age 0-4 years, but we could not see any change over time. Other risk factors remained similar **(Supplementary Table E2 and E3)**.

## Discussion

This study of childhood cancer survivors treated with lung-toxic treatments found that half of the population had at least one abnormal pulmonary function parameter at their last follow-up. FEV1 was below the lower limit of normal (LLN) in 33%, FVC in 38%, TLC in 22% and DLCO in 25%. There was no evidence for a significant change of FEV1 and FVC over time within a median observation period of 5.5 years. FEV1 and FVC were lower in survivors treated with thoracic surgery and did not improve over time, while FEV1 in children exposed to lung-toxic chemotherapy tended to improve during the observation period.

### Strengths and limitations

Strengths of this study are the large, national multi-center cohort of relatively unselected survivors including all types of childhood cancer subgroups and exposure to at least one lung-toxic treatment. The analysis of longitudinal data on lung function outcomes measured according to currently recommended protocols, and the long follow-up time after diagnosis strengthen the analyses. Further we used internationally recognized, reliable lung function outcomes and recommended reference equations to calculate percent predicted values and z-scores. Limitations included the retrospective nature of the study, which might have introduced selection bias of children presenting with respiratory symptoms. The included survivors might not be representative for all survivors exposed to lung-toxic treatment but reflect rather those with a higher exposure, more late effects, or those with respiratory symptoms. Though we used reference equations from the global lung function initiative, these might not perfectly fit the population in Switzerland.

### Prevalence of abnormal PFT measurements

In this cohort, 49% of included survivors had one or more pulmonary function outcomes below the LLN. In an independent Swiss study including 17 survivors exposed to lung toxic treatment, proportions with FEV1 and FVC below LLN were similar (43). The proportion of survivors with at least one condition (31%) was slightly lower than in a longitudinal study from the US by Armenian et al with similar, but slightly more conservative, cut-off values for obstructive disease, restrictive disease and DLCO impairment (46%), a single center US study by Green et al with an older patient population (65%) and a Dutch study by Mulder et al (44%) (11, 39, 44). Obstructive disease, defined as FEV1/FVC<0.7 and FEV1<80% of predicted value was rare in all studies (our cohort: <1%, Armenian: 4%, Green: 0.8%, Mulder: 2%) (11, 39, 44). Restrictive disease, defined as a TLC below the LLN was similar in our cohort (22%) and in the study by Armenian (18%) and Mulder (24%) but lower than reported by Green et al (49.9%). The population of Green et al was older when studied (34 years) than ours (17 years). Restrictive disease after childhood cancer might develop late in life and cancer treatment protocols in the 1990’s contained higher doses of radiotherapy and chemotherapy than nowadays. DLCO impairment, defined as <75% of predicted value, was less common in our cohort (18%), than in the study by Armenian et al (35%) and Green et al (45%). Screening for pulmonary dysfunction can detect many asymptomatic patients – Landier et al showed that pulmonary dysfunction was newly identified in 84% of at-risk patients (45). In Switzerland the screening mostly consists of spirometry and bodyplethysmography, survivors are less often referred for DLCO testing (46).

### Longitudinal trajectories

In our study, FEV1 and FVC trajectories remained below expected values throughout the entire follow-up period with no evidence for change over time. Considering that our population was rather young and heterogeneous in terms of the underlying cancer diagnosis and treatment, and included only the first 10 years after diagnosis, it is possible that deterioration may become more pronounced with longer follow-up. Until now longitudinal data on lung function of survivors was only studied for specific cancer subgroups such as embryonal brain tumors, where lung function (FEV1 and FVC) show similar trajectories as we found (17). In studies on HSCT survivors (19, 20), and survivors treated with thoracic irradiation for Hodgkin’s lymphoma (21), pulmonary function initially decreased, followed by a partial recovery and a subsequent slow decrease.

### Risk factors

In our study, patients with thoracic surgery had lower FEV1 and FVC z-scores than those without and remained low over time. In contrast, children exposed to lung-toxic chemotherapy started at a slightly lower FEV1 z-score but improved over time. As most participants had more than one exposure, the effect of individual lung-toxic treatment modalities might be underestimated and for some exposures such as HSCT and lung toxic chemotherapy we lack statistical power. As a result, poor lung function in children treated with surgery might reflect results of multimodal treatment strategies and not surgery alone **(Supplementary Table E4)**. Inclusion criteria and risk factors differed between other studies and ours, so that results are difficult to compare. Other studies found negative associations between female sex and diffusion impairment and FEV1 and FVC (11, 19). We could not find any effect of sex on lung function or lung function change over time. In a sensitivity analysis we found that survivors with older age at diagnosis tended to have a lower FEV1. This might indicate potential catch up growth in those younger at time of treatment (47). One study found that younger age at radiotherapy was associated with pulmonary dysfunction. However, the strength of the associations in this study diminished when adjusted for time since treatment (48).

### Implications

Pulmonary function surveillance may enable timely intervention potentially preventing unfavorable respiratory disease outcomes. Treatment options exist, for example, in individuals with bronchiolitis obliterans following HSCT (49).

Inconsistent use of reference and cutoff values on pulmonary dysfunction in previous studies make interpretation of the results and international comparison difficult. Also, the use of z-scores and LLN instead of percentage predicted affects results to some extend as we have shown (**Table 2**). We believe that it is time to agree on standardized reference and cutoff values to make research on pulmonary disease in survivors more homogenous and comparable. The reference equations provided by the GLI could be a valid option (27). Spirometry and body plethysmography are most commonly used to follow up survivors but may not be sensitive enough to detect subtle changes occurring in the lung periphery. Therefore, two recent studies used inert gas washout techniques in survivors (43, 50). Both studies could show that the test is feasible in survivors, even in younger children. However, the inert gas washout is only available in highly specialized clinics and the clinical impact has to be further investigated.

In conclusion the large proportion of survivors with reduced lung function identified in this study underlines the need for long-term surveillance of this vulnerable population and for prospective studies of lung toxicity after treatment for cancer in childhood.

## Data Availability

All data produced in the present study are available upon reasonable request to the authors

## Abbreviations

CNS: Central nervous system
CSI: Craniospinal irradiation
CI: Confidence interval
DLCO: Diffusion capacity of the lung for carbon monoxide
FEV1: Forced expiration volume in the first second
FVC: Forced vital capacity
HSCT: Hematopoietic stem cell transplantation
ICCC-3: International Classification of Childhood Cancer, Third edition
IQR: Interquartile range
LLN: Lower limit of normal
PFT: Pulmonary function tests
SCCR: Swiss Childhood Cancer Registry
SPOG: Swiss Pediatric Oncology Group
TLC: Total lung capacity

## Acknowledgements

We thank all childhood cancer survivors and families for participating in the SCCR. We thank the data managers of the SPOG (Claudia Althaus, Brigitte Andratschke, Nadine Assbichler, Pamela Balestra, Heike Baumeler, Nadine Beusch, Sarah Blanc, Susann Drerup, Yves Franzosi, Janine Garibay, Franziska Hochreutener, Sabine Holzapfel, Monika Imbach, Friedgard Julmy, Eléna Lemmel, Rodolfo Lo Piccolo, Heike Markiewicz, Veneranda Mattielo, Annette Reinberg, Marion Rizo, Renate Siegenthaler, Astrid Schiltknecht, Sophie Steiner, Beate Schwenke, and Verena Stahel) and the data managers and administrative staff of the SCCR (Meltem Altun, Erika Brantschen, Katharina Flandera, Anna Glenck, Elisabeth Kiraly, Eleftheria Michalopolou, Erika Minder, Verena Pfeiffer, Shelagh Redmond, and Cornelia Stadter).

## Supporting Information

**Supplementary Explanation E1** Explanation of multivariable linear mixed regression analysis with random intercept and random slope

**Supplementary Table E1** Lung function in childhood cancer survivors exposed to lung-toxic therapies on the last test when all five outcomes were measured by z-scores and percentage predicted at the same time (N=128)

**Supplementary Table E2** FEV1 z-scores in childhood cancer survivors exposed to pulmotoxic treatment compared to GLI 2012 reference values, secondary analysis including age at cancer diagnosis; multivariable linear mixed regression adjusted for all covariates in the model (180 subjects, 760 tests)

**Supplementary Table E3** FVC z-scores in childhood cancer survivors exposed to pulmotoxic treatment compared to GLI 2012 reference values secondary analysis including age at cancer diagnosis; multivariable linear mixed regression adjusted for all covariates in the model (178 subjects, 758 tests)

**Supplementary Table E4** Description of childhood cancer survivors treated with thoracic surgery, N=19

**Supplementary Figure E1** Flow chart of study population

