## Supplemental Material for "Longitudinal assessment of lung function in Swiss childhood cancer survivors"

**Supplementary Explanation E1** Explanation of multivariable linear mixed regression analysis with random intercept and random slope

The multivariable linear mixed regression analysis with random intercept and random slope allows to take clustering and longitudinal changes of tests from each patient into account.

Here we explain the data output with the example of FEV1:

The upper part of the table describes the FEV1 z-scores at time "zero", which refers to the time at diagnosis. It can be considered as the extrapolated baseline FEV1 z-score, where each patient starts at diagnosis. The reference patient (female, with lymphoma, no radiotherapy, no chemotherapy, no HSCT and no thoracic surgery) starts at a z-score of -1.07. If a person is male (z-score estimate 0.16) and additionally received thoracic radiotherapy (z-score estimate 0.16), he starts at a modelled FEV1 z-score of -0.75 (-1.07 (baseline) + 0.16 (difference female to male) + 0.16 (difference from no radiotherapy to radiotherapy)).

The lower part of the table describes the change of FEV1 z-scores per year. The reference patient (female, with lymphoma, no radiotherapy, no chemotherapy, no HSCT and no thoracic surgery) has an annual increase in FEV1 z-score of 0.09. For a male person (z-score estimate -0.07) exposed to thoracic radiotherapy (z-score estimate -0.05) the annual change in FEV1 z-score would be -0.03.

With this approach the starting point (intercept) and annual decrease (slope) can be calculated for patients with different diagnoses and exposure to different lung toxic treatment modalities.

**Supplementary Table E1** Lung function in childhood cancer survivors exposed to lung-toxic therapies on the last test when all five outcomes were measured by z-scores and percentage predicted at the same time (N=128)

| | Survivors | | |
|---|---|---|---|
| | n | % | median (IQR) |
| **Time from diagnosis to last test with all five outcome parameters** | 127 | | 5.0 (3.1 – 7.0) |
| **Age at last PFT (median, IQR [years])** | 127 | | 16.8 (12.4 – 19.2) |
| **FEV1** | | | |
| FEV1 z-score | 127 | | -1.0 (-2.0 – -0.1) |
| FEV1 z-score <-1.645 | 41 | 32 | -2.5 (-3.2 – -2.0) |
| FEV1 % predicted | 127 | | 88.4 (76.8 – 98.7) |
| FEV1 <80%predicted | 38 | 30 | 68.7 (61.3 – 74.8) |
| **FVC** | | | |
| FVC z-score | 127 | | -1.1 (-2.1 – -0.2) |
| FVC z-score <-1.645 | 47 | 37 | -2.6 (-3.3 – -2.0) |
| FVC % predicted | 127 | | 87.1 (76.0 – 98.1) |
| FVC <80% predicted | 44 | 35 | 68.5 (62.3 – 76.1) |
| **FEV1/FVC** | | | |
| FEV1/FVC z-score | 127 | | 0.2 (-0.4 – 1.1) |
| FEV1/FVC z-score <-1.645 | 5 | 4 | -2.3 (-2.6 – -1.9) |
| FEV1/FVC % predicted | 127 | | 101.0 (97.2 – 106.7) |
| FEV1/FVC <0.7 | 3 | 2 | 78.2 (67.7 – 79.9) |
| FEV1/FVC <0.7 and FEV1<80%predicted | 2 | 2 | n.a. |
| FEV1/FVC <0.7 and FEV1 z-score <-1.645 | 3 | 2 | n.a. |
| **TLC** | | | |
| TLC z-score | 127 | | -0.6 (-1.4– 0.04) |
| TLC z-score <-1.645 | 24 | 19 | -2.2 (-3.1 – -1.9) |
| TLC % predicted | 127 | | 92.6 (81.8 – 100.5) |
| TLC <75% predicted | 18 | 14 | 67.6 (50.2 – 73.2) |
| TLC <75% predicted and FEV1 ≥ 80% predicted | 2 | 2 | n.a. |
| **DLCO** | | | |
| DLCO z-score | 127 | | -0.7 (-1.5 – 0.5) |
| DLCO z-score <-1.645 | 29 | 23 | -2.4 (-3.4 – -1.8) |
| DLCO % predicted | 127 | | 89.7 (78.9 – 106.8) |
| DLCO <75% predicted[c] | 17 | 14 | 60.1 (53.5 – 69.2) |
| **FEV1, FVC, TLC or DLCO z-score < -1.645** | 63 | 50 | n.a. |

Abbreviations: IQR, interquartile range; PFT, pulmonary function test

**Supplementary Table E2** FEV1 z-scores in childhood cancer survivors exposed to pulmotoxic treatment compared to GLI 2012 reference values, secondary analysis including age at cancer diagnosis; multivariable linear mixed regression adjusted for all covariates in the model (180 subjects, 760 tests)

| | Estimate | 95% CI | | p-value * |
|---|---|---|---|---|
| **Intercept (FEV1 z-score at time of diagnosis †)** | -0.13 | -1.57 | 1.30 | |
| **Sex (ref. female)** | 0.09 | -0.38 | 0.56 | 0.707 |
| **BMI (ref. healthy: z-score ≥ -2, ≤ 1)** | | | | 0.458 |
| Underweight (z-score < -2) | -0.3 | -0.79 | 0.09 | |
| Overweight (z-score ≥ 1, ≤ 2) | 0.05 | -0.43 | 0.52 | |
| Obese (z-score > 2) | 0.10 | -0.54 | 0.73 | |
| **Cancer diagnosis (ref. lymphoma)** | | | | 0.119 |
| CNS tumor | -0.001 | -0.77 | 0.76 | |
| Bone tumors | -1.30 | -2.55 | -0.06 | |
| Germ cell tumors | 1.27 | -0.05 | 2.60 | |
| Leukemia | -0.77 | -1.97 | 0.43 | |
| Neuroblastoma | -1.39 | -3.27 | 0.49 | |
| Renal tumors | 0.25 | -1.52 | 1.03 | |
| Soft tissue sarcoma | -0.80 | -1.98 | 0.38 | |
| **Age at diagnosis (ref. 0 - 4 years)** | | | | |
| 5 - 13 years | -0.81 | -1.71 | 0.10 | 0.004 |
| >13 years | -0.96 | -1.89 | -0.04 | |
| **Thoracic radiotherapy (ref. no)** | 0.13 | -0.87 | 1.14 | 0.794 |
| **Chemotherapy (ref. no)** | -0.39 | -0.92 | 0.14 | 0.150 |
| **HSCT (ref. no)** | 0.17 | -0.64 | 1.00 | 0.678 |
| **Thoracic surgery (ref. no)** | -1.04 | -1.98 | -0.11 | 0.029 |
| **Change of FEV1 z-score per year** | | | | |
| **Time since diagnosis (continuous, per year)** | 0.06 | -0.19 | 0.31 | |
| **Interaction male sex** | -0.07 | -0.15 | 0.01 | 0.103 |
| **Interaction BMI (ref. healthy: z-score ≥ -2, ≤ 1)** | | | | 0.953 |
| Underweight (z-score < -2) | 0.003 | -0.15 | 0.16 | |
| Overweight (z-score ≥ 1, ≤ 2) | 0.003 | -0.09 | 0.10 | |
| Obese (z-score > 2) | 0.04 | -0.10 | 0.17 | |
| **Interaction cancer diagnosis (ref. lymphoma)** | | | | 0.059 |
| CNS tumor | -0.07 | -0.20 | 0.06 | |
| Bone tumors | 0.16 | -0.05 | 0.36 | |
| Germ cell tumor | -0.27 | -0.51 | -0.05 | |
| Leukemia | 0.12 | -0.08 | 0.33 | |
| Neuroblastoma | 0.29 | -0.04 | 0.61 | |
| Renal tumors | -0.05 | -0.23 | 0.13 | |
| Soft tissue sarcoma | 0.01 | -0.20 | 0.19 | |
| **Age at diagnosis (ref. 0 - 4 years)** | | | | 0.047 |
| 5 - 13 years | -0.02 | -0.15 | 0.11 | |
| >13 years | 0.09 | -0.05 | 0.23 | |
| **Interaction thoracic radiotherapy (ref. no)** | -0.05 | -0.24 | 0.14 | 0.582 |
| **Interaction chemotherapy (ref. no)** | 0.11 | 0.01 | 0.20 | 0.027 |
| **Interaction HSCT (ref. no)** | -0.16 | -0.27 | 0.002 | 0.046 |
| **Interaction thoracic surgery (ref. no)** | -0.04 | -0.19 | 0.12 | 0.639 |

Abbreviations: CNS, central nervous system; HSCT, hematopoietic stem cell transplantation

* Likelihood ratio test p-value indicating whether the characteristic explains differences in $FEV_1$ within the study population.

† Reference patient corresponds to a female person, at diagnosis, diagnosed with lymphoma, and not exposed to thoracic radiotherapy, chemotherapy, HSCT, and thoracic surgery.

**Supplementary Table E3** FVC z-scores in childhood cancer survivors exposed to pulmotoxic treatment compared to GLI 2012 reference values secondary analysis including age at cancer diagnosis; multivariable linear mixed regression adjusted for all covariates in the model (178 subjects, 758 tests)

| | Estimate | 95% CI | | p-value*¶ |
|---|---|---|---|---|
| **Intercept (FVC z-score at time of diagnosis †)** | -1.0 | -2.38 | 0.37 | |
| **Sex (ref. female)** | 0.02 | -0.41 | 0.46 | 0.910 |
| **BMI (ref. healthy: z-score ≥ -2, ≤ 1)** | | | | 0.360 |
| Underweight (z-score < -2) | -0.20 | -0.58 | 0.18 | |
| Overweight (z-score ≥ 1, ≤ 2) | -0.23 | -0.66 | 0.21 | |
| Obese (z-score > 2) | 0.15 | -0.41 | 0.71 | |
| **Cancer diangosis (ref. lymphoma)** | | | | 0.402 |
| CNS tumor | -0.06 | -0.75 | 0.61 | |
| Bone tumors | -0.19 | -1.33 | 0.94 | |
| Germ cell tumors | 1.26 | -0.01 | 2.53 | |
| Leukemia | -0.12 | -1.22 | 0.98 | |
| Neuroblastoma | -1.04 | -2.70 | 0.62 | |
| Renal tumors | 0.17 | -0.97 | 1.31 | |
| Soft tissue sarcoma | -0.53 | -1.61 | 0.55 | |
| **Age at diagnosis (ref. 0 - 4 years)** | | | | 0.339 |
| 5 - 13 years | -0.47 | -1.26 | 0.33 | |
| >13 years | -0.61 | -1.43 | 0.20 | |
| **Thoracic radiotherapy (ref. no)** | 0.41 | -0.59 | 1.41 | 0.424 |
| **Chemotherapy (ref. no)** | 0.15 | -0.33 | 0.63 | 0.535 |
| **HSCT (ref. no)** | -0.45 | -1.20 | 0.29 | 0.232 |
| **Thoracic surgery (ref. no)** | -1.41 | -2.26 | -0.55 | 0.001 |
| **Change of FVC z-score per year** | | | | |
| **Time since diagnosis (continuous, per year)** | 0.12 | -0.08 | 0.33 | |
| **Interaction male sex** | -0.04 | -0.11 | 0.02 | 0.168 |
| **Interaction BMI (ref. healthy: z-score ≥ -2, ≤ 1)** | | | | 0.101 |
| Underweight (z-score < -2) | -0.06 | -0.19 | 0.07 | |
| Overweight (z-score ≥ 1, ≤ 2) | 0.09 | 0.01 | 0.18 | |
| Obese (z-score > 2) | 0.07 | -0.04 | 0.18 | |
| **Interaction cancer diagnosis (ref. lymphoma)** | | | | 0.126 |
| CNS tumor | -0.08 | -0.18 | 0.02 | |
| Bone tumors | -0.09 | -0.25 | 0.07 | |
| Germ cell tumor | -0.18 | -0.37 | -0.004 | |
| Leukemia | 0.01 | -0.16 | 0.17 | |
| Neuroblastoma | 0.17 | -0.09 | 0.42 | |
| Renal tumors | -0.13 | -0.27 | 0.0001 | |
| Soft tissue sarcoma | -0.06 | -0.20 | 0.09 | |
| **Age at diagnosis (ref. 0 - 4 years)** | | | | 0.046 |
| 5 - 13 years | -0.01 | -0.11 | 0.09 | |
| >13 years | 0.08 | -0.03 | 0.19 | |
| **Interaction thoracic radiotherapy (ref. no)** | -0.11 | -0.28 | 0.05 | 0.188 |
| **Interaction chemotherapy (ref. no)** | -0.01 | -0.08 | 0.06 | 0.758 |
| **Interaction HSCT (ref. no)** | -0.03 | -0.13 | 0.08 | 0.613 |
| **Interaction thoracic surgery (ref. no)** | 0.04 | -0.07 | 0.16 | 0.462 |

Abbreviations: CNS, central nervous system; HSCT, hematopoietic stem cell transplantation

* Likelihood ratio test p-value indicating whether the characteristic explains differences in $FEV_1$ within the study population.

† Reference patient corresponds to a female person, at diagnosis, diagnosed with lymphoma, and not exposed to thoracic radiotherapy, chemotherapy, HSCT, and thoracic surgery.

**Supplementary Table E4** Description of childhood cancer survivors treated with thoracic surgery, N=19

| | Diagnosis | Year of diagnosis | Age at first thoracic surgery [years] | Relapse | Description of thoracic surgery/ surgeries | Additional lung toxic exposure[1] |
|---|---|---|---|---|---|---|
| 1 | (VIa) Nephroblastoma | 1992 | 6.6 | Yes | Thoracotomy, resection upper lobe left side, resection lower lobe right side | Mediastinal radiotherapy |
| 2 | (IIb) Non-Hodgkin lymphoma | 1995 | 13.7 | No | Laminectomy Th2-Th5 | Craniospinal irradiation |
| 3 | (VIIIc) Ewing tumor | 1995 | 5.6 | Yes | Rib resection, resection of lingula | Whole lung irradiation |
| 4 | (IIb) Non-Hodgkin lymphoma | 1996 | 16.4 | Yes | Thoracotomy, en bloc resection upper lobe left side | Mediastinal radiotherapy, busulfan, autologous HSCT |
| 5 | (VIa) Nephroblastoma | 1998 | 6.3 | Yes | Thoracotomy, resection lower lobe right side | Whole lung irradiation, autologous HSCT |
| 6 | (Xc) Malignant gonadal germ cell tumors | 1999 | 15.3 | No | Thoracotomy, resection upper lobe left side | Mediastinal radiotherapy, bleomycin |
| 7 | (VIa) Nephroblastoma | 2000 | 2.3 | Yes | Thoracotomy, wedge resection | Whole lung irradiation |
| 8 | (VIIIc) Ewing tumor | 2001 | 10.0 | Yes | Rib resection 7-9, Thoracotomy for resection of metastasis | Whole lung irradiation |
| 9 | (IIa) Hodgkin lymphoma | 2001 | 15.1 | No | Thoracoscopy, biopsy | Mantle irradiation, bleomycin |
| 10 | (VIIIc) Ewing tumor | 2002 | 14.3 | No | Thoracotomy, resection lower lobe left side, metastasectomy | Whole lung irradiation, autologous HSCT |
| 11 | (VIIIc) Ewing tumor | 2003 | 13.5 | No | Rib resection (6$^{th}$ rib) | *Radiotherapy other* |
| 12 | (IXd) Other specified soft tissue sarcoma | 2003 | 8.3 | No | Thoracotomy, resection lower lobe left side, resection of pleura and part of the diaphragm | Whole lung irradiation |
| 13 | (VIIIc) Ewing tumor | 2004 | 4.9 | Yes | Rib resection (rib 9-10) | Chest irradiation, autologous HSCT |
| 14 | (VIIIc) Ewing tumor | 2004 | 13.9 | Yes | Rib resection (rib 3-9), partial resection of the diaphragm | Radiotherapy other, autologous HSCT |
| 15 | (IIa) Hodgkin lymphoma | 2006 | 16.8 | Yes | Thoracoscopy, biopsy | Mediastinal radiotherapy, autologous HSCT |
| 16 | (VIIIc) Ewing tumor | 2007 | 12.9 | Yes | Thoracotomy, tumor resection, rib resection (9$^{th}$ rib) | |
| 17 | (VIIIc) Ewing tumor | 2009 | 14.6 | Yes | Thoracotomy, wedge resection, rib resection (rib 8-9) | Whole lung irradiation, autologous HSCT |
| 18 | (IXd) Other specified soft tissue sarcoma | 2009 | 8.1 | No | Thoracotomy, metastasectomy | Whole lung irradiation |
| 19 | (IXa) Rhabdomyosarcoma | 2012 | 14.6 | Yes | Thoracotomy, tumor resection, partial rib resection (rib 3-5) | Radiotherapy other |

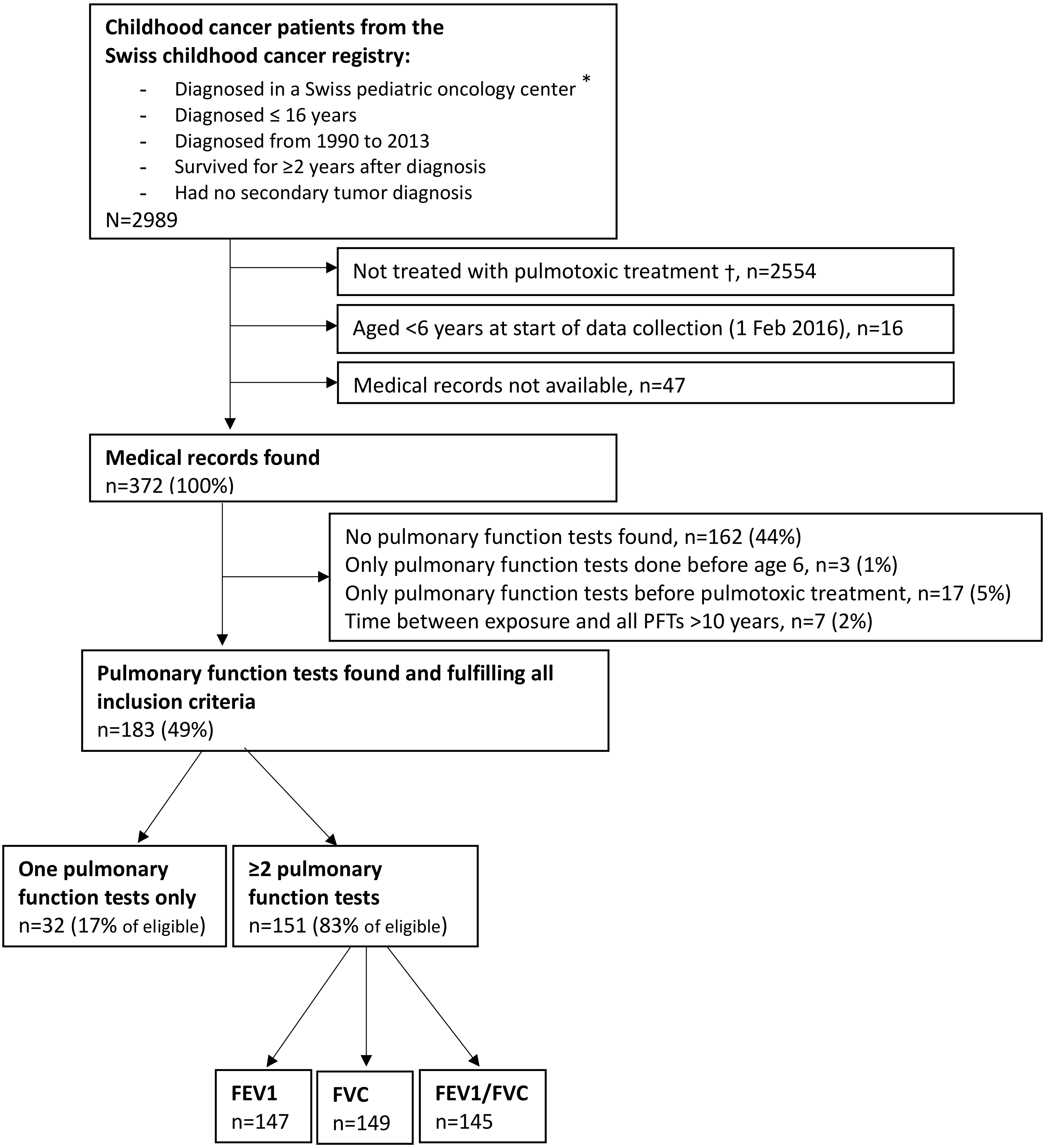

**Supplementary Figure E1** Flow chart of study population

*Including the following centers with pediatric oncology units: Kinderklinik Kantonsspital Aarau AG, Universitäts-Kinderspital Basel, Universitäts-Kinderklinik Inselspital Bern, Hospital des Enfants Geneve, CHUV Lausanne, Kinderklinik Kantonsspital Luzern, Ostschweizer Kinderspital St. Gallen, Universitäts-Kinderspital Zürich

† Pulmotoxic treatment defined as chemotherapy with busulfan, bleomycin, lomustine or carmustine and/or chest radiotherapy
